# A Multi-Agent clinical pre-consultation system for structuring noisy patient reported information into clinical reports and AI-ready data

**DOI:** 10.64898/2026.06.10.26355372

**Authors:** Mengyan Zhang, Jiyun Zhao, Wenjun Tang, Junjie Li, Jie Xing, Han Zhang, Junyao Qiu, Yan Zhang

## Abstract

In primary care and outpatient settings, clinically important patient information is often embedded in fragmented, ambiguous, repetitive, and noisy communication between physicians and patients. This limits physicians’ ability to obtain a clear pre-consultation overview of symptoms, history of present illness, and visit intent, while also preventing real world clinical dialogues from being reused in hospital information systems and medical artificial intelligence applications. To address this challenge, we developed PCRAgent, a centrally coordinated multi agent framework for pre-consultation clinical information organization, shifting information processing upstream from the consultation. Guided by physician inquiry logic, PCRAgent identifies, extracts, corrects, and standardizes patient-reported information from noisy consultations. Its coordinated modules including error detection, semantic editing, output control, contextual memory, and intent recognition enable robust parallel handling of spelling errors, repetitions, grammatical inconsistencies, medical ambiguities, and non-medical interference. A traceable edit list records intermediate corrections and context, allowing iterative refinement without redundant modifications. PCRAgent generates two complementary outputs. One is a Pre-Consultation Clinical Report for rapid physician review. The other is a Structured Clinical Conversation Dataset for hospital data construction and downstream AI applications. In evaluations using 220,000 strongly perturbed consultations, PCRAgent maintained high robustness, achieving a clinical information accuracy of 4.99 out of 5 and key element completeness of 5 out of 5, outperforming GPT4o. Expert review of Chinese and English dialogues confirmed high clinical accuracy of 4.85 out of 5 and high security of 4.79 out of 5. Multicenter validation in real world outpatient workflows further demonstrated practical utility. These results indicate that PCRAgent improves outpatient workflow efficiency, reduces physicians’ cognitive burden, ensures completeness of pre-consultation clinical information, supports more focused and accurate clinical decision-making, and enables high-quality reuse of clinical data for downstream medical artificial intelligence applications.

## Introduction

The rapid expansion of digital health platforms has generated large volumes of medical dialogues, creating new opportunities for automated electronic health record (EHR) generation, clinical decision support, and medical artificial intelligence[1, 2]. However, in primary care and outpatient settings, these dialogues are often fragmented, semantically imprecise, repetitive, emotionally expressive, or dialect-influenced, particularly among older adults and patients with communication difficulties. Clinicians must therefore spend consultation time clarifying patient statements, organizing symptom trajectories, and reconstructing the history of present illness before clinical decision-making. This communication burden reduces outpatient efficiency, increases the risk of incomplete information capture, and prevents real-world dialogue from being reliably converted into structured clinical evidence.

The problem becomes more pronounced when outpatient consultations are digitized. Clinical dialogue data are frequently contaminated by automatic speech recognition errors, informal expressions, abbreviations, irrelevant text, and context-dependent ambiguity [3, 4]. Such noise can propagate into downstream large language model (LLM) applications, where misrecognized medications, missing negations, or distorted symptoms may compromise the security and accuracy of generated clinical documentation [5]. Existing rule-based tools can correct simple surface errors but are too rigid for complex medical dialogue, whereas general-purpose large language models are prone to over-correction, semantic drift, and hallucination [6–9]. More fundamentally, these approaches are mainly designed for text correction or free-form summarization rather than for generating structured, traceable, and clinically usable pre-consultation records or standardized datasets suitable for downstream AI applications [10, 11].

To address these limitations, we propose PCRAgent, a multi-agent framework that converts noisy patient consultations into physician-ready pre-consultation records and reusable structured clinical data. Coordinated by a central controller, PCRAgent integrates four specialized modules, including Error Detector, Semantic Editing, Output Control, and Interactive modules, to detect communication errors, perform clinically constrained correction, verify output quality, and standardize consultation content. The framework extracts core clinical elements, including chief complaint, symptom characteristics, history of present illness, relevant medical history, and visit intent, and generates a Pre-Consultation Clinical Report for rapid physician review before the formal encounter. This report provides a structured overview of patient-reported symptoms, symptom trajectories, and consultation purpose, helping reduce repetitive clarification and support more focused clinical assessment. PCRAgent also produces a Structured Clinical Conversation Dataset for hospital data construction, clinical documentation, downstream medical AI applications, and future large language model training. By linking pre-consultation support with reusable clinical data generation, PCRAgent addresses both the clinical communication burden and the data-structuring bottleneck in real-world outpatient care.

## Results

### Overview of PCRAgent

To enable reliable generation of structured pre-consultation clinical records, it is essential to first understand whether noisy patient-reported clinical information can be directly standardized using existing large language models. We therefore conducted a preliminary evaluation to examine how real-world consultation noise affects downstream clinical understanding and response generation. In clinical environments, patient dialogues are frequently compromised by heterogeneous noise, including ASR errors, informal expressions, and irrelevant interruptions. To quantify this impact, we conducted a preliminary analysis in which GPT-4o processed perturbed consultations with increasing noise intensities. We observed that escalating noise levels substantially degraded consultation quality, as reflected by consistent declines in Accuracy, Integrity, Smoothness, and Semantic Similarity **(Fig. S1)**. Correspondingly, the generated replies increasingly failed to meet clinical standards, particularly in Completeness, Empathy, and Clarity, even though surface-level accuracy remained relatively stable. These results indicate that direct LLM-based processing is insufficient to reliably standardize noisy patient-reported clinical information. The failure arises from the lack of explicit error localization, traceable correction mechanisms, and structured output verification, which are essential for preserving clinically meaningful content under heterogeneous noise conditions. This limitation motivates the need for a dedicated system-level solution capable of robust clinical information organization.

PCRAgent adopts a collaborative multi-agent architecture guided by a Parallel Feedback Strategy initiated by a central scheduler. Within this framework, the system decomposes the task into four specialized modules. The Error Detector serves as the analysis layer and identifies multiple types of noise in medical dialogues in parallel. The Semantic Editing module performs targeted corrections as the execution layer. A traceable EditList is maintained as structured working memory to store edit traces and contextual states. The Output Control module continuously evaluates output quality and triggers iterative refinement when predefined criteria are not satisfied. Finally, the Interactive module organizes the corrected information into structured clinical outputs. This mechanism ensures transparent, verifiable, and controlled transformation of noisy patient-reported information while reducing redundant modifications **(Fig. 1a-b)**.

**Figure 1.**
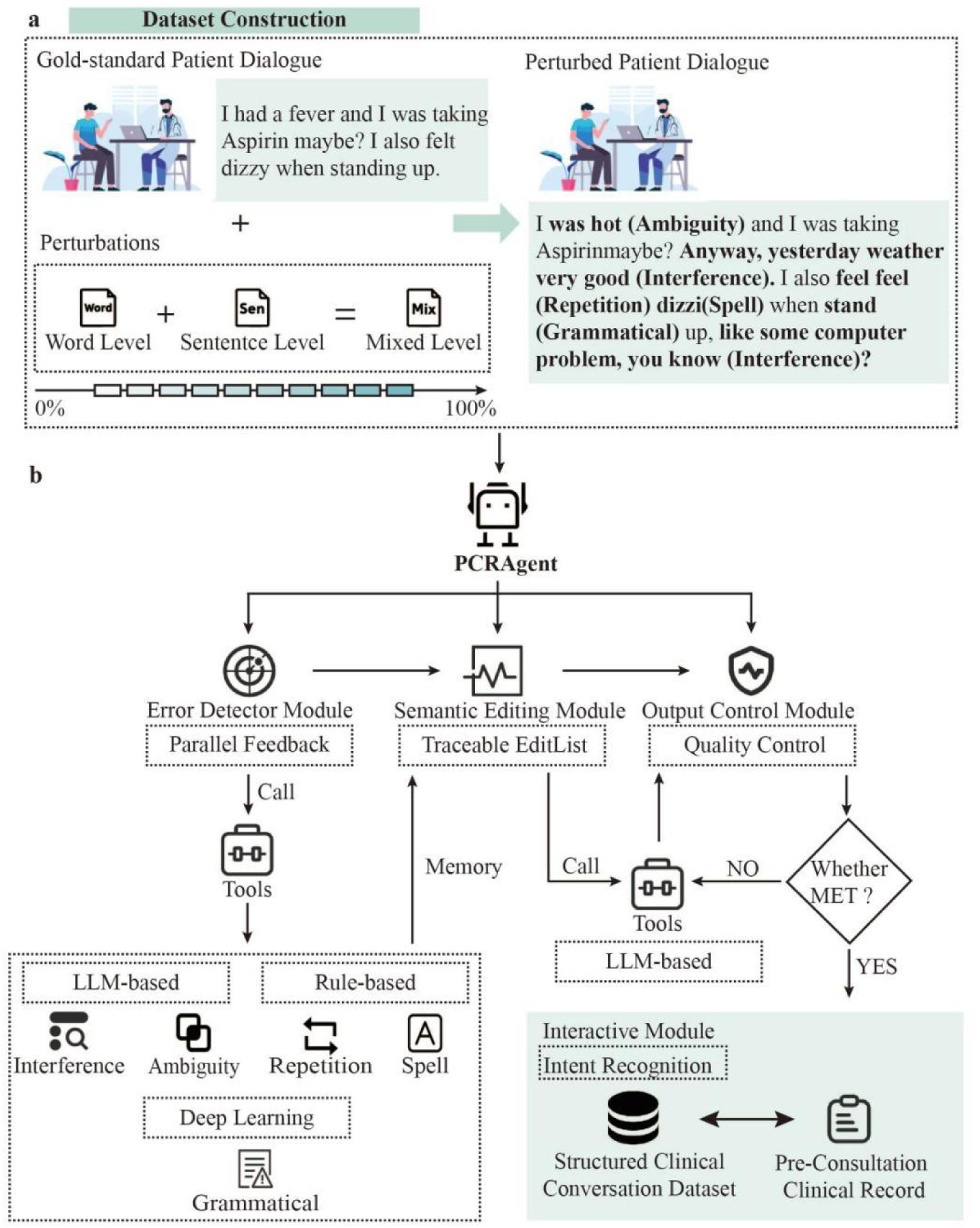
Overview of PCRAgent. **(a)** Five types of noise are injected into gold-standard patient dialogues to create perturbed patient dialogues. **(b)** A central scheduler decomposes the task to four modules: Error Detector (calls tools to identify five noise types in parallel), Semantic Editing (performs targeted corrections. The five identified error types are stored as memory into the traceable EditList), Output Control (evaluates quality and triggers re-planning via a traceable EditList), and Interactive Module (generates professional responses). This produces two outputs: a Structured Clinical Conversation Dataset and a Pre-Consultation Clinical Report (PCR).

PCRAgent operates through a structured multi-stage workflow that transforms noisy patient-reported clinical information into standardized outputs suitable for both physician review and downstream data reuse **(Fig. S2a-c)**. The process begins with consultation semantic restoration via the Error Detector module. Given a noisy input, the module performs parallel analysis across five categories of clinical noise, including spelling errors, repetitions, grammatical inconsistencies, medical ambiguities, and non-medical interference. To preserve clinical validity, medical terminology is protected through constraint-aware detection supported by a medical knowledge graph, preventing clinically meaningful terms such as drug names and disease entities from being incorrectly modified. Rather than directly rewriting the consultation, the module produces structured error annotations that explicitly localize and categorize noise, enabling fine-grained downstream correction.

Based on these annotations, the Semantic Editing module performs targeted restoration of patient-reported information. Instead of full-text rewriting, only annotated regions are modified, and all edits are recorded in a traceable EditList. This design enables memory-guided refinement, allowing unresolved or partially corrected regions to be revisited in subsequent iterations while preserving already validated clinical content.

The standardized patient-reported information is then passed to the Output Control module for session-level quality verification **(Fig. S2b)**. This module evaluates whether the edited consultation satisfies predefined criteria for Accuracy, Integrity, and Smoothness. When deficiencies are detected, only the corresponding regions are selectively refined using the EditList, ensuring global consistency without redundant reprocessing of the entire consultation. This iterative verification loop continues until clinically acceptable standardization is achieved.

Following verification, the Interactive module performs high-level semantic synthesis of the standardized patient-reported information and inferred consultation intent **(Fig. S2c)**. Based on this synthesis, the system generates a Pre-Consultation Clinical Report, which organizes key clinical elements including chief complaint, history of present illness, symptom characteristics, and patient intent into a physician-oriented format. In parallel, the standardized interaction data are structured into a Structured Clinical Conversation Dataset for hospital information systems and downstream medical artificial intelligence applications.

Together, this workflow enables PCRAgent to convert fragmented and noisy patient communication into clinically structured outputs through coordinated detection, editing, verification, and synthesis stages. The resulting Pre-Consultation Clinical Report provides physicians with rapid pre-encounter understanding of patient condition, while the Structured Clinical Conversation Dataset supports scalable clinical data reuse and infrastructure development.

### PCRAgent maintains robust session-level standardization of patient-reported clinical information under increasing dialogue noise

We next evaluated whether PCRAgent can maintain stable session-level standardization performance under increasing levels of clinical dialogue noise. Each complete interaction was treated as one pre-consultation session, where physician questions serve as clinical inquiry anchors and patient responses provide the patient-reported information to be extracted, corrected, and organized. The resulting session-level standardized output was evaluated using Accuracy, Integrity, Smoothness, and Semantic Similarity, which together measure clinical meaning preservation, information completeness, readability, and semantic consistency.

Across noise intensities ranging from 10 percent to 100 percent, PCRAgent maintained consistently high standardization quality **(Fig. 2a-d)**. At 10 percent noise, the standardized session-level output achieved an Accuracy of 4.99 (SD 0.12) and an Integrity of 4.97 (SD 0.26). Performance showed no cumulative degradation as noise increased. Even at 100 percent noise, Accuracy remained 4.99 (SD 0.07) and Integrity reached 5.00 (SD 0.00). These results indicate that PCRAgent can reliably preserve patient-reported clinical information even under dense and heterogeneous noise conditions.

**Figure 2.**
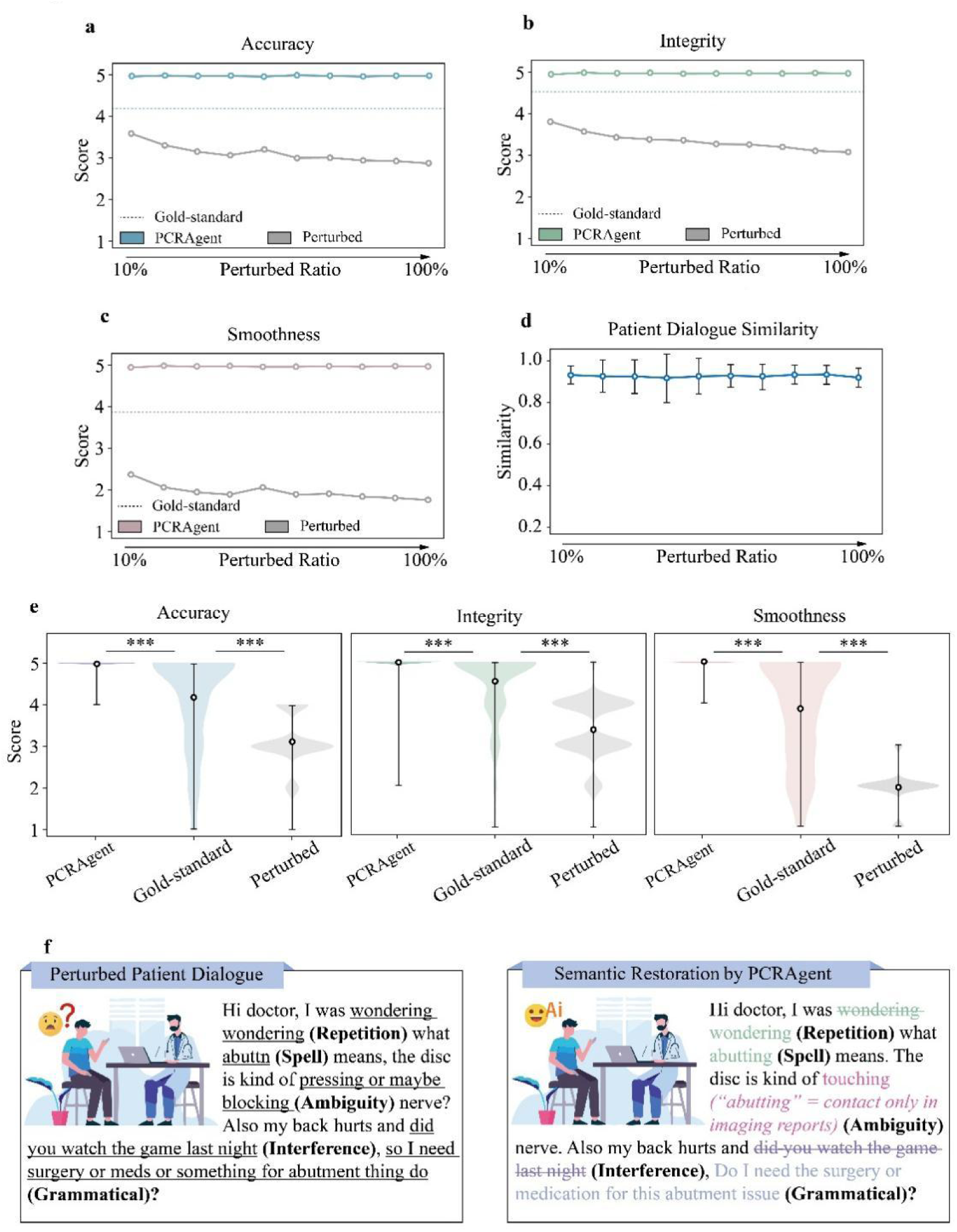
Consultation-level restoration performance of PCRAgent under increasing noise. **(a–d)** Consultation-level restoration quality evaluated at perturbation ratios ranging from 0% to 100%, measured by Accuracy, Integrity, Smoothness, and Semantic Similarity, respectively. Scores are computed on the fully restored consultation text after PCRAgent processing, reflecting the combined effect of error detection, semantic editing, and output control across all error types. **(e)** Statistical comparison of overall restoration performance among three groups: perturbed consultations, PCRAgent consultations, and Gold-standard consultations, with all perturbation levels aggregated. **(f)** A representative example of a noisy clinical consultation and its corresponding restored version generated by PCRAgent, illustrating targeted correction of multiple error types while preserving valid medical content.

This robustness arises from the coordinated effect of the Parallel Feedback strategy and the Output Control module. Parallel Feedback enables simultaneous detection of multiple error types, reducing cascading error propagation across the session. The Output Control module enforces session-level consistency by selectively refining unresolved regions based on previously stored correction traces, rather than regenerating the entire consultation. As a result, corrections remain localized while global clinical consistency is preserved.

We further compared PCRAgent with rule-based methods (LanguageTool and SymSpell), a fine-tuned sequence-to-sequence model (T5), and a general-purpose large language model (GPT-4o) using the same evaluation metrics **(Fig. 3a-b)**. PCRAgent consistently outperformed all baselines in standardization quality while maintaining stable computational efficiency. Rule-based methods showed substantial degradation under heterogeneous noise, reflecting limited clinical language coverage. GPT-4o preserved fluency but exhibited reduced clinical fidelity under extreme noise, indicating that direct generation may introduce semantic drift and omission of clinically relevant information.

**Figure 3.**
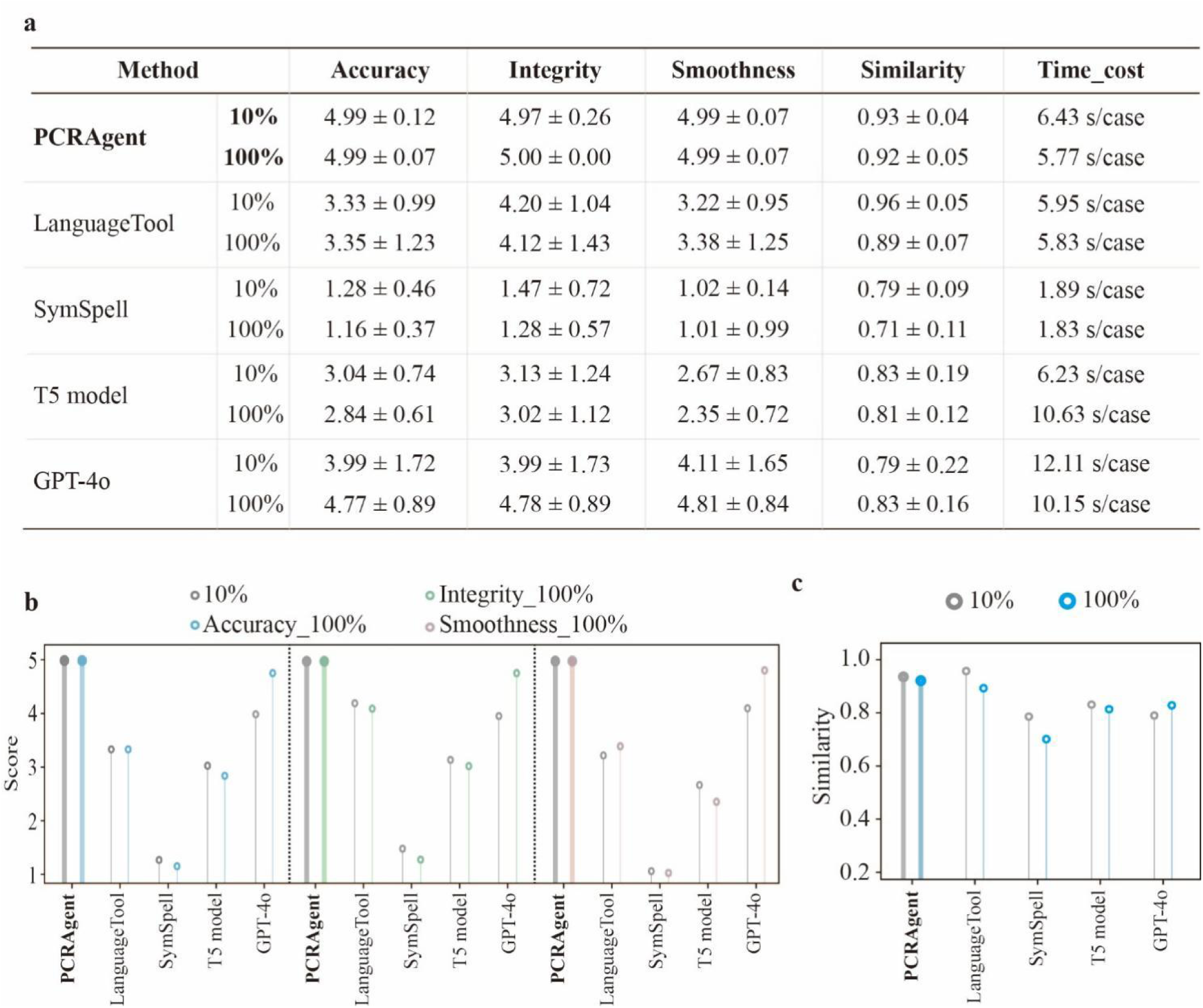
Comparison with baseline methods. **(a)** Quantitative comparison of consultation-level restoration quality across PCRAgent and baseline methods, including rule-based approaches (LanguageTool, SymSpell), a fine-tuned sequence-to-sequence model (T5), and a full LLM (GPT-4o). All methods are evaluated on fully restored consultation text using Accuracy, Integrity, Smoothness, and Semantic Similarity under the same noise conditions. **(b)** Bar chart comparing restoration quality across different methods. Restoration quality is evaluated using three metrics: Accuracy, Integrity, and Smoothness, which respectively measure factual correctness, completeness of preserved information, and linguistic fluency after semantic editing. **(c)** Bar chart comparing semantic similarity between different methods at two perturbation levels (10% and 100%) on consultation-level restoration.

In terms of efficiency, PCRAgent required 6.43 seconds per session at 10 percent noise compared with 12.11 seconds for GPT-4o, and remained stable at 5.77 seconds at 100 percent noise. This reflects the advantage of annotation-guided and memory-guided refinement, which avoids redundant full-text regeneration.

We further evaluated generalization using bilingual clinical dialogue datasets **(Fig. S4a-b)**. PCRAgent achieved comparable performance across English and Chinese consultations, with no statistically significant differences in Accuracy, Integrity, or Smoothness. Although Semantic Similarity was lower in Chinese, Accuracy and Integrity remained stable, suggesting that clinically relevant meaning was preserved across language settings.

Overall, these results demonstrate that PCRAgent can reliably standardize patient-reported clinical information under increasing noise levels, across baseline comparisons, and across languages. This session-level standardized representation serves as a stable intermediate layer for downstream generation of Pre-Consultation Clinical Reports and Structured Clinical Conversation Datasets.

### Ablation analysis confirms the contribution of specialized detectors to robust session-level standardization

To determine which components supported the robust performance of PCRAgent, we performed detector-level ablation within the Error Detector module. Each specialized detector was removed independently, including spelling, grammar, repetition, medical ambiguity, and non-medical interference detectors. Standardized session-level consultation outputs were evaluated at 10 percent and 100 percent noise using Accuracy, Integrity, Smoothness, and Semantic Similarity **(Fig. 4a-c)**.

**Figure 4.**
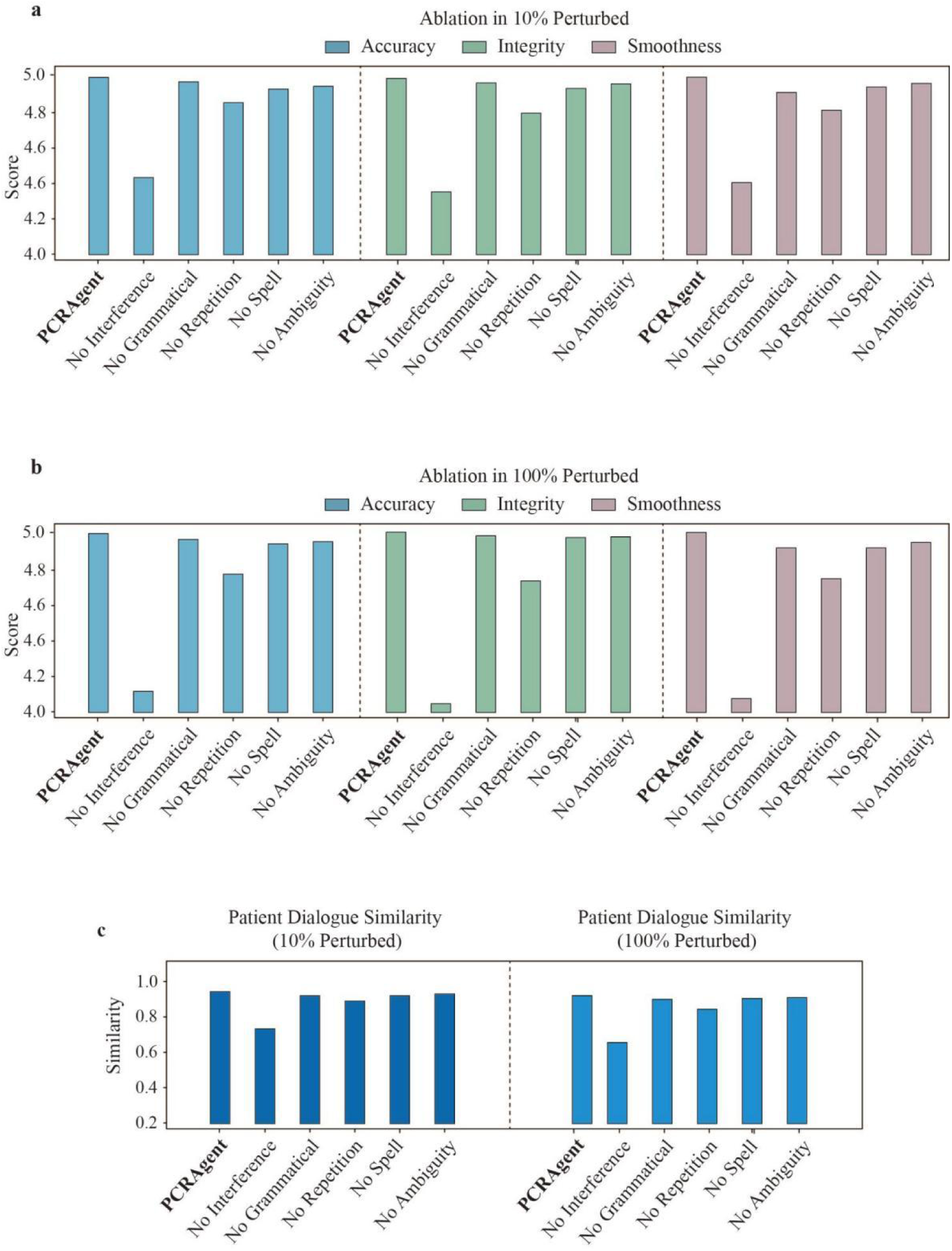
Contribution of parallel error detectors within the Error Detector Module. **(a)** Ablation results on consultation texts with 10% perturbation. Each specialized detector (spelling, grammar, repetition, medical ambiguity, or non-medical interference) is removed independently, and restoration quality is evaluated using Accuracy, Integrity, and Smoothness. **(b)** Ablation results on consultation texts with 100% perturbation, evaluated using the same metrics as in (a), illustrating detector contributions under extreme noise conditions. **(c)** Semantic Similarity between restored consultations and reference text for both 10% and 100% perturbation settings, comparing the full Error Detector Module with individual detector removals.

At 10 percent noise, the complete Error Detector module achieved the highest performance, with Accuracy of 4.99, Integrity of 4.97, and Smoothness of 4.99 **(Fig. 4a)**. Removing spelling or grammar detection caused only modest declines, indicating that these detectors mainly support local fluency and readability. Removal of medical ambiguity detection moderately reduced Accuracy and Integrity, reflecting its role in preserving clinically relevant meaning. In contrast, removing repetition detection caused broader performance loss, while removal of non-medical interference detection produced the largest decline, reducing Accuracy, Integrity, and Smoothness to 4.43, 4.35, and 4.40, respectively.

At 100 percent noise, this hierarchy became more evident **(Fig. 4b)**. The full module maintained high Accuracy, Integrity, and Smoothness, whereas removal of repetition detection reduced these scores to 4.77, 4.73, and 4.74. Removal of non-medical interference detection caused the greatest degradation, with Accuracy decreasing to 4.12, Integrity to 4.05, and Smoothness to 4.08. These findings indicate that repeated expressions and irrelevant conversational content become major sources of standardization failure under dense and heterogeneous noise. Semantic Similarity followed the same pattern **(Fig. 4c)**. The complete module maintained high Semantic Similarity at both 10 percent and 100 percent noise. Removal of spelling or grammar detection produced mild changes, whereas removing repetition or non-medical interference detection substantially reduced Semantic Similarity, particularly under extreme noise. This suggests that repetition and irrelevant content affect not only readability but also the preservation of global consultation meaning. These results demonstrate that parallel detection of heterogeneous noise is essential for robust session-level standardization. Different detectors contribute at distinct levels, with spelling and grammar detectors improving local text quality, medical ambiguity detection preserving clinical meaning, and repetition and non-medical interference detection maintaining global coherence and clinical relevance. This multi-detector design provides a reliable standardized input for downstream Pre-Consultation Clinical Report generation and Structured Clinical Conversation Dataset construction.

### The Output Control module is required for reliable session-level standardization of patient-reported clinical information

To evaluate whether local error detection and semantic editing alone are sufficient for robust patient-reported information standardization, we assessed the contribution of the Output Control module within PCRAgent. We compared the full framework with an ablated variant that retained only the Error Detector and Semantic Editing modules. This setting evaluates whether local correction alone can ensure globally consistent session-level clinical information under realistic pre-consultation conditions, where physician questions provide clinical anchors and patient responses contain noisy clinical content to be standardized.

Across all noise levels, removing the Output Control module resulted in substantial degradation in Accuracy, Integrity, Smoothness, and Semantic Similarity **(Fig. 5a-d)**. At 10 percent noise, the ablated variant achieved an Accuracy of 2.65 (SD 0.90), an Integrity of 2.24 (SD 1.28), and a Smoothness of 2.43 (SD 0.94). These results indicate that although local error detection and targeted semantic editing can correct isolated issues, they are insufficient to maintain globally coherent and clinically complete session-level patient-reported information.

**Figure 5.**
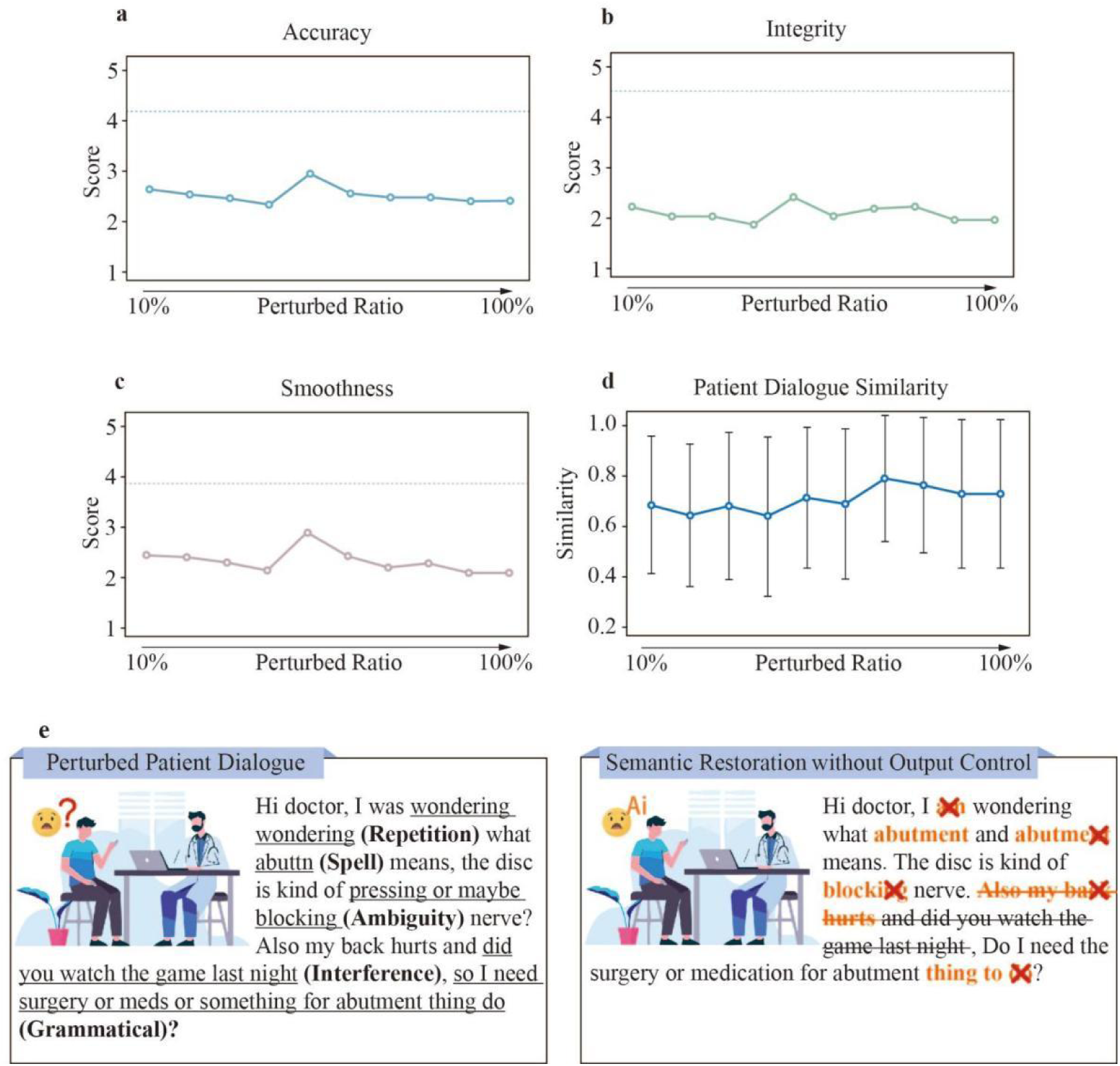
Contribution of the Output Control Module to consultation-level restoration. **(a–d)** Comparison of restoration quality across perturbation ratios ranging from 0% to 100%, evaluated using Accuracy, Integrity, Smoothness, and Semantic Similarity, respectively. **(e)** A representative example illustrating the output generated without the Output Control Module.

The performance degradation became more severe as noise increased. At 100 percent noise, Accuracy decreased to 2.42 (SD 0.77) and Integrity to 1.98 (SD 1.06), with Smoothness and Semantic Similarity showing consistent decline. These findings suggest that without output-level verification, residual and partially corrected errors accumulate across the session, leading to loss of global coherence, incomplete information reconstruction, and reduced clinical readability under dense and heterogeneous noise.

In contrast, the full PCRAgent framework maintains stable session-level standardization through the Output Control module. This module acts as a quality gate that evaluates whether the standardized patient-reported information satisfies predefined criteria for Accuracy, Integrity, and Smoothness. When deficiencies are detected, it retrieves structured correction history from the EditList and selectively refines unresolved regions. This memory-guided verification prevents redundant full-session rewriting, preserves validated clinical content, and enforces global consistency after local semantic correction.

Together, these results demonstrate that Output Control is essential for transforming locally corrected patient narratives into globally consistent session-level standardized clinical information. By introducing system-level verification on top of error detection and semantic editing, PCRAgent ensures that the standardized output remains reliable for downstream generation of Pre-Consultation Clinical Reports and construction of Structured Clinical Conversation Datasets.

### PCRAgent-standardized patient dialogues supports intent recognition and downstream clinical downstream utility

To evaluate the downstream utility of PCRAgent-standardized patient reported clinical information, we conducted a controlled clinical language generation experiment using three input conditions, including perturbed patient reported information, PCRAgent standardized information, and gold-standard information. Across all perturbation levels, PCRAgent-standardized inputs consistently produced higher-quality clinical outputs than perturbed inputs and approached gold-standard performance across Accuracy, Completeness, Safety, Empathy, and Clarity **(Fig. 6a)**. Aggregated analysis further confirmed statistically significant improvements over perturbed inputs across all evaluation dimensions (Mann-Whitney U test, P < 0.001; **Fig. 6b**). These results indicate that structured patient-reported information improves the reliability of downstream clinical language generation.

**Figure 6.**
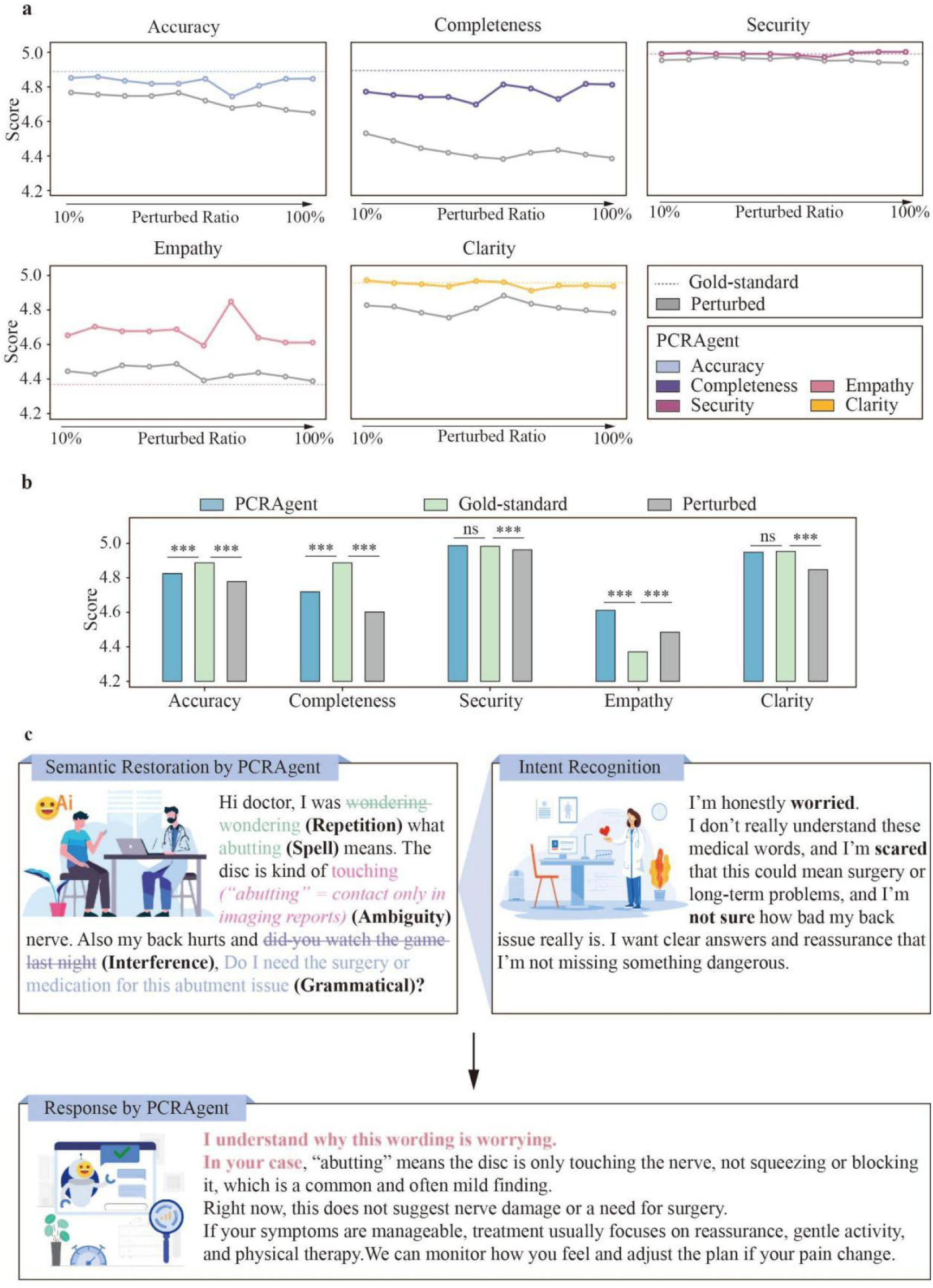
Downstream utility evaluation based on PCRAgent-standardized patient dialogue. **(a)** Evaluation of downstream clinical responses across perturbation ratios from 10 percent to 100 percent using Accuracy, Completeness, Security, Empathy, and Clarity. Responses were generated by the same LLM using three input conditions, including perturbed patient dialogue, PCRAgent-standardized patient dialogue, and gold-standard patient dialogue. **(b)** Aggregated statistical comparison of response quality among the three input conditions across all perturbation levels. Statistical significance was assessed using the Mann-Whitney U test. **(c)** Representative example showing how PCRAgent restores noisy patient-reported information and reconstructs patient intent before downstream response generation. PCRAgent corrected spelling, ambiguity, interference, and grammatical errors, clarified the patient concern, and supported a response that addressed both the clinical content and the patient emotional context.

A representative case further illustrates the role of intent-aware standardization **(Fig. 6c)**. PCRAgent not only restores noisy patient expressions but also identifies incomplete or ambiguous clinical information and reconstructs underlying patient intent. In this example, the system clarifies patient concerns related to nerve injury risk, surgical intervention, and long-term prognosis. Importantly, this intent reconstruction is derived from explicit patient expressions and contextual clinical cues, enabling more structured and patient-centered downstream responses.

This behavior reflects an additional function of PCRAgent beyond noise correction. By identifying missing or implicit clinical information during the standardization process, the intent recognition module facilitates more complete and structured patient-reported outputs. This contributes to more informative clinical representations, which can further improve downstream language generation performance.

Overall, these findings suggest that PCRAgent standardized patient-reported clinical information enhances the usability of noisy clinical dialogue data by improving both semantic clarity and intent completeness. This supports its role as a pre-consultation clinical intelligence layer that provides structured inputs for Pre-Consultation Clinical Report generation, Structured Clinical Conversation Dataset construction, and downstream medical AI applications.

### Real-world validation of PCRAgent-generated Pre-Consultation Clinical Reports

After demonstrating that PCRAgent can generate stable session-level standardized patient-reported clinical information, we evaluated its final physician-facing pre-consultation clinical report, in real world outpatient settings. Three independent clinical experts assessed each report using a 5-point Likert scale across Accuracy, Completeness, Clinical Safety, and Clarity. Inter-rater reliability was high, with an intraclass correlation coefficient greater than 0.85, indicating consistent evaluation across experts.

We validated PCRAgent using 35 real-world outpatient dialogues collected from two geographically distinct medical centers in China, including Harbin Medical University (n = 18, Northern China) and Hainan Medical University (n = 17, Southern China) in May 2026. Across all cases, PCRAgent-generated reports achieved mean scores of 4.29 for Accuracy, 4.57 for Completeness, 4.83 for Clinical Safety, and 4.66 for Clarity **(Fig. 7a)**. No significant difference was observed between the two regional cohorts (P > 0.05; **Fig. 7b**), suggesting that PCRAgent maintains stable performance across heterogeneous clinical communication styles and institutional settings.

**Figure 7.**
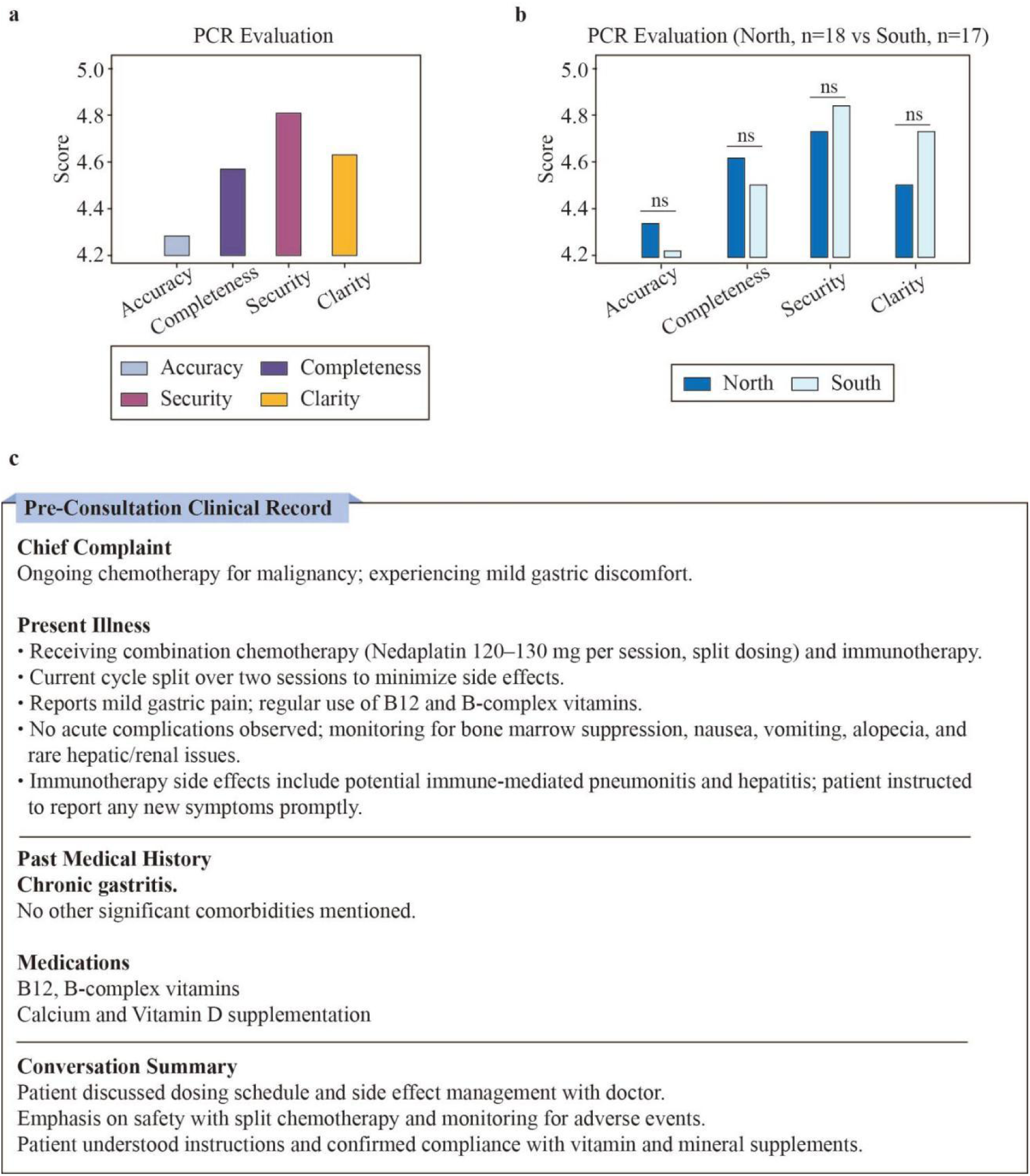
Evaluation of PCR generation quality. **(a)** PCR performance scores across four metrics under real-world outpatient dialogues. **(b)** Cross-regional generalizability comparison between the Southern cohort (Hainan Medical University, n = 17) and Northern cohort (Harbin Medical University, n = 18). **(c)** Demonstration of the detailed clinical content and structured components within a generated PCR.

A representative example illustrates how PCRAgent organizes standardized patient-reported information into structured clinical sections, including chief complaint, history of present illness, relevant medical history, and a consolidated summary incorporating patient intent and consultation concerns **(Fig. 7c)**. This structured format enables rapid physician review before the encounter and reduces the need for repeated clarification during consultation.

In parallel, the standardized patient-reported information can be systematically accumulated into a Structured Clinical Conversation Dataset, supporting hospital data integration and downstream medical artificial intelligence applications. Together, these findings demonstrate that PCRAgent provides a robust and generalizable framework for transforming noisy real-world patient communication into clinically usable pre-consultation documentation and reusable structured clinical data.

## Discussion

PCRAgent addresses an important communication and data structuring challenge in digital healthcare by transforming noisy patient reported information into two clinically meaningful outputs, a physician-facing Pre-Consultation Clinical Report and a reusable Structured Clinical Conversation Dataset. In primary care and outpatient settings, clinically relevant information is often embedded in fragmented, ambiguous, repetitive, dialect-influenced, or transcription-corrupted patient expressions. This makes it difficult to obtain a clear pre-encounter overview of the chief complaint, symptom trajectory, history of present illness, and visit intent. It also prevents real-world clinical dialogue from being reliably reused in hospital information systems and downstream medical AI applications. Prior studies have similarly shown that transcription errors and noisy dialogue can compromise clinical understanding and documentation pipelines in real-world settings [12]. By organizing noisy patient communication into structured pre-consultation documentation and standardized dialogue data, PCRAgent provides a practical bridge between real-world patient expression, physician workflow, and AI-ready clinical data infrastructure.

A central contribution of PCRAgent is that it treats noisy patient communication as a session-level clinical information organization problem rather than a generic text correction or summarization task. Direct rewriting by general-purpose large language models can improve fluency, but it may also introduce semantic drift, over-correction, omission of clinically relevant details, or hallucinated content when inputs contain dense and heterogeneous noise. PCRAgent instead decomposes the workflow into coordinated modules for error detection, semantic editing, output control, contextual memory, and intent recognition. This modular design allows the framework to identify different sources of noise, preserve valid medical terminology, correct targeted regions, and verify whether the standardized patient-reported information remains clinically accurate, complete, and readable.

The Parallel Feedback Strategy is central to this robustness. Rather than processing errors sequentially, PCRAgent detects spelling errors, grammatical inconsistencies, repeated content, medical ambiguities, and non-medical interference in parallel. This reduces the risk of cascading failures, in which errors missed at an early stage propagate into later outputs, a known weakness of sequential clinical text processing pipelines [13]. The ablation results indicate that different detectors contribute at distinct levels. Spelling and grammar detectors mainly improve local readability, whereas medical ambiguity detection helps preserve clinical meaning. Repetition and non-medical interference detectors are especially important for maintaining global coherence and clinical relevance, because unresolved redundancy or irrelevant content can distort the overall patient narrative. These findings suggest that robust standardization of patient-reported information requires coordinated multi-signal detection rather than generic rewriting alone, consistent with prior work emphasizing explicit multi-tool coordination and agentic planning in complex language processing tasks [14]. Another important feature of PCRAgent is its traceable editing workflow. The EditList records each correction, contextual state, and unresolved issue, creating an auditable intermediate representation between the original noisy input and the final standardized output. This differs from opaque full-text generation, where it is difficult to determine which parts of the input were modified, preserved, or omitted. In clinical settings, such traceability is valuable because trust depends not only on the quality of the final output, but also on whether the transformation process can be inspected and verified. The EditList also enables efficient memory-guided refinement, allowing unresolved regions to be revisited without repeatedly regenerating the entire session. This helps preserve already validated clinical content and reduces unnecessary modification of stable information. The use of structured working memory for state-aware reasoning has also been recognized as an important mechanism for improving explainability and targeted refinement in complex AI workflows[15].

The Output Control module further distinguishes PCRAgent from systems that rely only on detection and editing. Local correction alone is insufficient to ensure that a complete pre-consultation session remains accurate, complete, and coherent. The Output Control module acts as a session-level quality gate by evaluating whether the standardized patient-reported clinical information satisfies predefined criteria for accuracy, integrity, and readability. When deficiencies remain, the module retrieves relevant correction history from the EditList and selectively refines unresolved regions. This closed-loop verification explains why PCRAgent maintained stable performance under increasing noise levels and why the full framework substantially outperformed ablated versions without output control.

The clinical value of PCRAgent lies in its ability to convert standardized patient-reported information into a Pre-Consultation Clinical Report. This report organizes core elements such as chief complaint, history of present illness, symptom characteristics, relevant medical history, patient intent, and core consultation concerns into a physician-oriented format. Such a structure can help physicians rapidly review the patient condition and visit purpose before the formal encounter, reduce repetitive clarification, and support more focused clinical assessment. Importantly, this role should be understood as pre-consultation information organization rather than autonomous diagnosis. PCRAgent does not replace physician judgment. Instead, it prepares patient-reported information in a clearer and more structured form so that clinical encounters can begin from a more complete information baseline.

In parallel, PCRAgent generates a Structured Clinical Conversation Dataset that supports the data infrastructure needs of digital healthcare. Real-world clinical dialogues are often difficult to reuse because they are noisy, inconsistent, and poorly structured. By converting session-level patient-reported information into standardized records with traceable fields and intent annotations, PCRAgent enables clinical dialogue data to be accumulated, governed, and reused for hospital databases, clinical documentation workflows, quality control, and future medical AI development. This dual-output design is important because the same workflow supports both immediate physician-facing utility and long-term data reuse.

The downstream clinical language generation experiment should be interpreted as an auxiliary utility test rather than as a core clinical function of PCRAgent. Its purpose was to determine whether PCRAgent standardized patient-reported clinical information could serve as a more reliable input for downstream medical language tasks. Responses generated from PCRAgent standardized inputs were more accurate, complete, safe, empathetic, and clear than those generated from perturbed inputs, and in some dimensions approached gold-standard performance. These findings suggest that PCRAgent preserves clinically relevant content and clarifies patient intent in a way that improves downstream model usability. This is consistent with prior robustness studies showing that targeted noise handling can substantially improve output quality relative to generic models [16]. PCRAgent is also notable for its operational feasibility. The framework maintained strong performance without requiring specialized hardware, indicating that robust consultation standardization, PCR generation, and structured data construction can be achieved under modest computational conditions. This feature is especially relevant for resource-constrained healthcare settings, where the practical adoption of AI systems often depends not only on model performance but also on deployability. The observed balance between quality, latency, and memory demand suggests that PCRAgent has value not only as a methodological contribution but also as a workflow-oriented system with realistic implementation potential.

Several limitations remain. First, although PCRAgent performed robustly under the evaluated perturbation settings, it may still face challenges when encountering unseen terminology, newly introduced drug names, emerging clinical slang, rare disease-specific expressions, or institution-specific abbreviations. Second, PCR generation currently relies primarily on information available within the standardized pre-consultation interaction and does not yet incorporate dynamic external resources such as drug databases, or institution-specific knowledge bases. This may partly explain residual gaps relative to gold-standard inputs in some downstream tasks. Future development should focus on knowledge integration, retrieval from clinical guidelines, drug resources, institutional protocols, and specialty-specific terminology banks may improve adaptation to evolving clinical language and domain-specific requirements. In conclusion, PCRAgent provides a clinically oriented framework for transforming noisy patient-reported communication into physician-ready pre-consultation reports and AI-ready structured clinical data resources.

## Methods

### Dataset Construction

All clinical dialogue samples in this study are drawn from the AI Medical Chatbot dataset, a publicly available corpus containing patient queries paired with detailed physician recommendations. We selected the first 1,273 dialogues as the experimental set, providing a representative range of patient expressions, symptom descriptions, and medical explanations for constructing controlled noisy conditions.

To simulate realistic disruptions in clinical environments, we implemented a mixed-level perturbation framework combining sentence-level interference and word-level perturbations. Sentence-level interference models environmental interruptions, such as background conversations or overlapping speech. For each consultation, a non-medical sentence sampled from MultiWOZ 2.2—a large-scale task-oriented dialogue dataset covering hotel booking, transport, and other everyday topics—is inserted at a random position. The inserted segment ranges from 10–100% of the original sentence length, reflecting scenarios where medical statements are interrupted mid-sentence, requiring the system to recover semantic content while filtering irrelevant information.

Word-level perturbations introduce finer-grained distortions, including spelling errors, grammatical inconsistencies, ambiguous tokens, and repeated fragments. Corrupted tokens are inserted into each sentence, with the total noise amount scaled to 10–100% of the original character count. The error-type proportions in distortion—27.2% spelling, 35.8% grammar, 18.4% ambiguity, and 18.6% repetition—are derived from an empirical analysis of 35 publicly available English medical dialogue videos from YouTube OSCE Guide, UKMLA, and CPSA materials. These audio materials were manually transcribed, and the resulting statistics **(Fig. S3)** provide an empirical basis for simulating realistic conversational noise.By combining sentence- and word-level perturbations, the final dataset captures both structural interruptions and token-level corruption. This mixed-level design creates a controlled yet challenging evaluation environment, enabling assessment of PCRAgent’s ability to detect errors, preserve medical terminology, and restore coherent clinical intent under complex noisy conditions.

To further evaluate the agent’s clinical usability under authentic clinical variations, we prospective constructed a dedicated Real-World Consultation Dataset during May 2026, comprising 35 genuine Chinese clinical consultation cases. These real-world data were multi-centrically collected from the outpatient departments of two geographically distinct institutions in China: 17 cases from Hainan Medical University (representing Southern China) and 18 cases from Harbin Medical University (representing Northern China). To guarantee clinical diversity, the cases specifically targeted outpatients from the Department of Cardiovascular Medicine and the Department of Dermatology. Evaluation of this dataset enables a rigorous performance comparison between simulated perturbation benchmarks and authentic clinical PCR outcomes.

### Unified evaluation framework for patient reported clinical information standardization and clinical output assessment

All evaluations in this study are conducted using DeepSeek-V3 under a unified and fixed scoring rubric. The same evaluator is applied both within the Output Control Module of PCRAgent for internal quality assessment and during final result evaluation, ensuring consistency across all stages. Evaluation is implemented following the GEval framework[17], which has been widely adopted for structured LLM-based assessment of generated medical text.

For consultation standardization quality, three dimensions are assessed: Accuracy, Integrity, and Smoothness. Accuracy measures resistance to hallucination and factual deviation introduced during consultation standardization. Integrity captures the extent to which clinically essential information from the original consultation is preserved without omission or distortion. Smoothness evaluates linguistic fluency and readability after editing, focusing on whether the standardized consultation remains natural and coherent **(Table S2)**.

For recommendation responses generated from standardized consultations, five complementary dimensions are evaluated: Accuracy, Completeness, Security, Empathy, and Clarity. Accuracy verifies that medical statements remain aligned with established clinical knowledge and do not introduce hallucinated content. Completeness assesses whether all clinically relevant aspects of the patient’s concerns are adequately addressed. Security screens for subtle Security risks, such as premature reassurance, speculative interpretation, or misleading framing. Empathy evaluates whether the response meaningfully incorporates psychological intent inferred from the patient’s subtext, rather than relying on generic or template-based comforting language. Clarity measures whether the structure and expression of the recommendation allow patients to easily understand the explanation and guidance provided **(Table S3)**.

All metrics are rated on a 5-point Likert scale, where 1 indicates poor performance and 5 indicates high performance. The selection of these evaluation dimensions is informed by prior studies on clinical dialogue assessment and Securityoriented medical text evaluation[18]. To further ensure clinical relevance and interpretability, two physicians with over five years of outpatient clinical experience reviewed the candidate metrics and finalized the evaluation protocol.

Within the Output Control Module of PCRAgent, fixed acceptance thresholds are applied to regulate output quality. These thresholds are derived from the mean scores assigned by DeepSeek-V3 to the gold-standard consultation dataset using the same evaluation rubric, and therefore serve as a calibrated reference for acceptable performance. Specifically, outputs are required to satisfy Accuracy ≥ 4.2, Integrity ≥ 4.5, and Smoothness ≥ 3.9, ensuring that only standardized consultation outputs meeting gold-standard-level quality are released.

To bridge the gap between automated metrics and real-world clinical utility, we established a rigorous human evaluation protocol specifically for the Pre-Consultation Clinical Report (PCR). The expert panel consisted of three senior clinicians from the Affiliated Hospital of Harbin Medical University, each possessing extensive outpatient experience and high academic standing in their respective fields: a Chief Physician in General Medicine with 10 years of clinical practice, a Deputy Chief Physician in Cardiology with 8 years of experience, and a Senior Consultant in Respiratory Medicine with 10 years of experience. This diverse panel ensured that the PCRs were evaluated against the stringent standards of actual clinical workflows.

The expert evaluation was conducted using a double-blind, randomized protocol to eliminate potential biases. The panel assessed the PCRs across four dimensions essential for clinical decision support: Accuracy, Completeness, Security, and Clarity. To ensure the reliability of the scores, the Inter-Rater Reliability (IRR) was calculated using the Intraclass Correlation Coefficient (ICC).

### Error Detector Module for Parallel Clinical Noise Detection

The Error Detector Module identifies and encodes all noise in perturbed consultation text—including spelling mistakes, repetitions, grammatical inconsistencies, medical ambiguities, and non-medical interference—while preserving the original sequence. Following the Parallel Feedback strategy, five detectors run simultaneously, each dedicated to a single error type. Their outputs are merged into a single list of structured annotations, where each annotation contains five fields: start_char, end_char, op, candidate_texts, and confidence. These annotations provide a consistent and minimal format for subsequent semantic restoration in the Semantic Editing Module.

### Medical Term Protection Mechanism

To prevent clinically important terms from being incorrectly flagged, the module embeds a Medical Term Protection Mechanism implemented via the MedicalTermsManager. This manager loads a JSON lexicon derived from Webster’s New World Medical Dictionary (Third Edition) and stores entries in a constant-time lookup set. Optional prefix checks are supported by a Trie, but detection primarily relies on hashed membership tests for efficiency.

During detection, each token is first passed to MedicalTermsManager.is_medical_term(token). Tokens present in the lexicon are exempt from all noise detection—including spelling mistakes, repetitions, grammatical inconsistencies, medical ambiguities, and interference—ensuring valid medical terminology remains untouched. Tokens absent from the lexicon proceed to standard detection procedures. The lexicon is comprehensive for established medical terminology, including drug names, symptoms, and disease labels, but may not cover emerging clinical slang or newly introduced drugs. Future work will integrate dynamic knowledge retrieval to supplement the static lexicon, allowing detection of zero-shot terms.

Parallel Feedback StrategyDetection is decomposed into independent channels operating simultaneously on the same input. Each detector focuses on a predefined error type, producing structured annotations in a uniform format. Although detectors operate in parallel, potential conflicts—such as overlapping annotations from spelling and grammatical detectors—are resolved downstream by the Output Control Module, which fine-tunes the standardized consultation output after semantic editing. This design ensures that multiple feedback signals contribute coherently to subsequent processing without introducing inconsistencies.

For spelling mistakes, tokens are first checked by a deterministic SpellChecker against a predefined dictionary. Tokens that fail validation are passed to SymSpell, which computes Damerau–Levenshtein distances with a threshold of 2. Detected errors produce SPL-type annotations with a REPLACE operation, 1–3 candidate corrections, and a confidence score derived from edit distance.

For repeated phrases, a sliding-window n-gram comparison (n = 1–3) is applied. Exact matches between adjacent n-grams generate DELETE-type annotations with a fixed confidence of 0.9.

For grammatical inconsistencies, the GECTagger generates a corrected sequence hypothesis. Token-level alignment between the original text and the hypothesis produces GRM-type annotations (REPLACE, INSERT, DELETE) without rewriting the input. Confidence scores are assigned based on edit distance and operation type. For medical ambiguities and non-medical interference, a lightweight LLM is employed as a constrained annotator, guided by a structured system prompt. The prompt defines the agent role and task, instructing it to (1) identify semantically ambiguous words in a medical context—including medical polysemy, conflicts between medical and common meanings, and ambiguous medical abbreviations—and (2) detect non-medical conversation fragments such as small talk, environmental noise, or unrelated technical topics. The prompt specifies strict output rules: ambiguous tokens are annotated as [AMB:original_word], non-medical fragments are enclosed within [ITF:start]…[ITF:end], and the original text must remain unchanged. All LLM outputs are parsed into structured annotations that integrate seamlessly with the parallel feedback pipeline.

All detectors produce annotations independently, forming a multi-layered feedback signal. Because all outputs follow a uniform schema and are non-destructive, they can be merged by the Output Control Module into a coherent set of annotations to guide Semantic Editing and support accurate, reliable consultation standardization. Semantic Editing Module for Traceable Consultation Standardization

The Semantic Editing Module processes all Edit entries produced by the Error Detector Module, including spelling errors (SPL), repeated phrases (RPT), grammatical errors (GRM), ambiguous terms (AMB), and non-medical interference (ITF), and converts them into a traceable, deterministic, and semantically coherent standardized consultation text. Each Edit retains its origin, type, detector source, and confidence score to ensure full traceability and reproducibility.To merge parallel edits, the module applies a structured procedure guided by a compatibility matrix. Compatible edit pairs, including GRM+SPL, RPT+GRM, and RPT+SPL, are merged by consolidating candidate texts, averaging confidence scores, and combining tags into a single merged Edit. Incompatible edits, particularly those involving AMB or ITF, are preserved independently to retain complete information. Edits are first sorted by starting position and then compared pairwise. Overlapping edits are merged when compatible and retained separately when not. Candidate texts are deduplicated and reordered to provide concise and relevant alternatives. The merged operation type is determined according to predefined rules, where DELETE+DELETE yields DELETE, INSERT+INSERT yields INSERT, and mixed operations yield REPLACE. Confidence scores for merged edits are computed using a weighted scheme that reflects relative edit types. For AMB and ITF entries, a lightweight LLM (GPT-4o-mini) is used as a constrained, non-generative annotator. For AMB entries, the model produces concise, context-aware English medical interpretations in a word(interpretation) format that remains directly linked to the original Edit for traceability. For ITF entries, the model confirms the annotation as non-medical and applies the corresponding DELETE operation. The system prompt enforces strict constraints to avoid rewriting or altering the original consultation text.After processing, all entries, including merged SPL, RPT, GRM, interpreted AMB, and confirmed ITF, are applied in reverse positional order to prevent index shifts. Only DELETE, REPLACE, and INSERT operations are executed. The final output consists of the processed EditList, the deterministic edit actions applied, the fully standardized consultation text, and the interpreted text in which all ambiguous terms have been resolved.This module preserves the parallel feedback mechanism, maintains semantic consistency across heterogeneous error types, and provides the standardized textual basis for downstream Output Control, PCR generation, and construction of the Structured Clinical Conversation Dataset.

Output Control Module for Conflict Resolution and Quality-Controlled Standardization. The Output Control Module serves as the final quality-control layer of PCRAgent, consolidating all edits produced by the Semantic Editing Module and ensuring that the standardized consultation output remains coherent and clinically accurate. A primary function of this module is to resolve positional conflicts between overlapping edits and to validate semantic correctness across the full consultation text. To perform this step, the module uses a lightweight LLM (GPT-4o-mini) as a constrained evaluator that jointly considers three complementary inputs. These include the full standardized EditList, the edited consultation text produced by the Semantic Editing Module, and the original perturbed consultation text. By comparing these three representations, the module can cross-validate deterministic corrections, semantically interpreted edits, and the untouched consultation content, thereby reducing reliance on any single potentially distorted representation.

Based on this joint view, the module selectively refines local regions that do not satisfy predefined quality criteria, while preserving validated content that already meets the required standard. This design enables conflict resolution, semantic consistency checking, and quality-controlled consultation standardization without restarting the full editing process from scratch. As a result, the Output Control Module provides the final standardized consultation text used for downstream Interactive processing, Pre-Consultation Clinical Report (PCR) generation, and construction of the Structured Clinical Conversation Dataset.

Rather than relying on fixed priority rules, GPT-4o-mini performs a semantic comparison across the three inputs by examining how each conflicting edit appears in both the original perturbed consultation text and the edited consultation text produced by the Semantic Editing Module. This comparison allows the model to determine whether competing edits convey the same clinical meaning, whether either edit contains medically relevant information, and whether the two proposed modifications can coexist in the final output.

Internally, the model reconstructs a localized semantic field around each conflicting region, evaluating neighboring tokens, their syntactic roles, and the communicative intent implied by surrounding dialogue. Through this reconstruction, GPT-4o-mini distinguishes several types of conflict: (1) edits imposing incompatible interpretations on the same clinical phrase, (2) edits addressing different but complementary aspects of the same expression—such as morphology correction versus ambiguity resolution, and (3) edits in the EditList that contradict the meaning already stabilized in the edited consultation text. By triangulating these signals across the three textual views, the model selects the most coherent resolution strategy, which may involve retaining a single edit, merging candidate rewrites when both provide valid corrections, or discarding and regenerating edits to better preserve clinical intent. Once conflicts are resolved, the module proceeds to quality assurance, reassessing the edited consultation text as a whole to ensure coherence within the full clinical context. This step is particularly critical for AMB annotations and ITF deletions. The model evaluates whether ambiguous terms are sufficiently clarified without altering clinical meaning, whether ITF elements are fully but not excessively removed, and whether any interaction among edits introduces unintended changes to symptom descriptions, temporal relations, or diagnostic cues. By leveraging all three textual views, the module identifies over-deletion by comparing the edited text to clinically relevant content preserved in the original consultation and detects under-editing by checking residual inconsistencies between the edited text and the candidate edits in the EditList.

### Output Control and Quality Assurance Module

After resolving all annotation-level conflicts, the Output Control Module performs a final layer of quality assurance to ensure that the consultation text meets clinical Security and communication standards. The module incorporates an automated Quality Evaluation Mechanism based on the GEval framework. Immediately after producing the final text, DeepSeek-V3 evaluates it along three calibrated dimensions: Accuracy, reflecting resistance to hallucination and factual deviation; Integrity, capturing preservation of clinically essential information; and Smoothness, measuring linguistic fluency. Fixed acceptance thresholds (Accuracy ≥ 4.2, Integrity ≥ 4.5, Smoothness ≥ 3.9) are derived from mean scores computed on the gold-standard patient-consultation dataset using the same evaluation rubric.

If the text fails any threshold, the module triggers the Self-Correction Algorithm. Rather than regenerating the entire sentence, the system constructs a structured feedback prompt that integrates three elements: the deficient evaluation dimensions, the original noisy input, and the EditList recording applied edits and their rationale. GPT-4o-mini then performs targeted revisions, selectively revisiting only the regions implicated by the failing dimensions. For example, a low Integrity score prompts re-examination of edits involving clinically salient entities or ITF deletions, while a low Smoothness score directs attention to grammar- and repetition-related edits. By grounding corrections in the provenance encoded in Edit objects, the system preserves traceable editing intent, avoids unnecessary full-sequence generation, and reduces semantic drift.

If the text still fails to meet thresholds after the maximum number of retries, a fallback selection policy is applied. All intermediate revisions are ranked hierarchically—Accuracy first, then Integrity, and finally Smoothness—and the highest-scoring version is selected as the final output. Through this layered evaluation and provenance-aware revision strategy, the Output Control Module ensures that the final consultation text is both clinically reliable and communicatively coherent.

### Interactive Module for Patient Intent-Aware Clinical Synthesis and Pre-Consultation Report Generation

The Interactive Module serves as the final synthesis layer of PCRAgent, transforming the standardized patient-reported clinical information into a structured Pre-Consultation Clinical Report (PCR). This module focuses on intent-aware clinical information organization and high-level semantic synthesis for physician review.

At its core, the module incorporates a lightweight intent recognition mechanism that captures patient-reported concerns and consultation intent from the standardized clinical text. To achieve this, the system first converts patient expressions into a first-person intent representation, enabling a more explicit modeling of underlying concerns that may not be directly stated in the dialogue. This representation is then mapped to a set of clinically and psychologically defined intent anchors, each describing a specific type of patient concern such as uncertainty about diagnosis, fear of disease progression, or need for reassurance **(Table S4)**. Both representations are encoded into embedding space, and intent alignment is performed through cosine similarity to identify the most relevant clinical intent category.

The inferred intent signal is integrated with the standardized patient-reported clinical information to form an intent-aware clinical representation. This representation serves as the input for structured clinical synthesis, enabling the system to organize key clinical elements into a Pre-Consultation Clinical Report. The report summarizes chief complaint, history of present illness, symptom characteristics, relevant medical history, and patient intent in a physician-oriented format, ensuring that both explicit clinical information and inferred consultation concerns are clearly represented.

In addition to structured report generation, the module performs semantic aggregation across the entire standardized consultation to produce a concise and clinically organized summary. This process reorganizes fragmented patient narratives into standardized clinical headers while preserving medically relevant terminology and ensuring readability. The resulting Pre-Consultation Clinical Report provides clinicians with a multi-dimensional overview of patient condition prior to consultation, supporting rapid pre-encounter understanding and reducing information fragmentation during clinical workflow.

Through intent representation learning, anchor based interpretation, and structured clinical synthesis, the Interactive Module ensures that patient-reported information is transformed into a coherent and clinically usable format.

### Baseline comparison experiments for semantic restoration

To contextualize PCRAgent’s performance, we evaluated a set of representative baseline systems spanning rule-based, fine-tuned, and general LLM paradigms. The comparison included LanguageTool, an open-source grammar correction engine; SymSpell, a high-efficiency rule-based spelling corrector; a fine-tuned T5 model, representing mainstream specialist architectures for English text restoration; and full LLM generation. All baselines were provided the same perturbed consultation texts and performed semantic restoration without access to internal editing signals.

We tested two perturbation levels—10% and 100%—corresponding to mild corruption and extreme semantic distortion. This design bypasses intermediate restoration steps, focusing on the system’s ability to handle both low and high noise scenarios. Restored outputs were evaluated along Accuracy, Integrity, and Smoothness, providing a multi-dimensional measure of restoration quality. Comparing performance across these perturbation extremes establishes baseline stability and quantifies the relative gains introduced by PCRAgent.

### LanguageTool for rule based grammatical correction

As a rule-based baseline, LanguageTool is applied as an off-the-shelf grammatical correction pipeline for noisy clinical dialogue, without task-specific customization. This ensures that restoration strictly follows predefined linguistic rules. In experiments, LanguageTool is configured with English (en-US) settings. Each noisy consultation utterance is processed independently: the system detects spelling errors, verb-form inconsistencies, article misuse, and punctuation issues, and applies corrections in a single pass using default mechanisms. No manual rule filtering, prioritization, or iterative refinement is introduced, and dialogue context, speaker roles, or turn-level information are ignored. The output is a grammatically corrected version of the original input. Semantic restoration accuracy is quantified using sentence-level similarity between the restored output and the gold-standard consultation text. Sentence embeddings are generated via a pretrained model, and cosine similarity measures alignment with intended clinical meaning rather than the noisy input. Evaluation follows the same framework as PCRAgent, including Accuracy, Integrity, and Smoothness to capture content correctness, completeness of medical information, and linguistic fluency. System efficiency is additionally assessed through processing time and token consumption, ensuring a fair comparison of computational overhead across methods.

### SymSpell for high efficiency spelling correction

SymSpell is applied as a high-efficiency, dictionary-based word-level spelling-correction pipeline, without any learning-based adaptation. It performs fast approximate string matching under strict edit-distance constraints. In experiments, SymSpell is initialized with a maximum edit distance of 2. A frequency-based English dictionary is loaded when available; otherwise, a lightweight internal dictionary of common English tokens ensures basic coverage. All entries are static and not updated during processing. Noisy consultation text is tokenized using regular-expression word boundaries while preserving punctuation and sentence structure. Each token is processed independently: candidate corrections are generated using SymSpell’s lookup mechanism, and the top match is selected based on dictionary frequency. Original capitalization is preserved. To prevent modification of clinical entities, a medical terminology lexicon is used to identify protected terms, which are copied directly to the output without alteration. The reconstructed sentence maintains original token order and punctuation. No sentence-level rewriting, grammatical restructuring, or context-aware adjustment is applied, and dialogue history or speaker information is ignored. Semantic alignment is quantified by computing sentence embeddings for the restored output and the gold-standard consultation text, with cosine similarity capturing recovery of the intended clinical meaning. Evaluation follows the same framework as PCRAgent, using Accuracy, Integrity, and Smoothness to measure content correctness, completeness of medical information, and linguistic fluency. Computational efficiency is additionally assessed via time cost and token consumption for direct comparison across methods.

The T5 model is applied as a fine-tuned sequence-to-sequence framework for text restoration. Noisy consultation text is prepended with the instruction “denoise:” to form the input prompt. The transformer-based encoder-decoder architecture encodes the full input sequence into contextualized representations, while the decoder autoregressively generates corrected tokens conditioned on the encoder output and prior tokens. The model and tokenizer are loaded via the HuggingFace transformers library using AutoTokenizer.from_pretrained and AutoModelForSeq2SeqLM.from_pretrained, with trust_remote_code=True to support custom architectures. Text generation uses the pipeline interface with do_sample=False for deterministic outputs and max_new_tokens=512 to constrain sequence length. Generated text is post-processed to remove the “denoise:” prefix and trim whitespace. Each input instance is processed independently. The model leverages pre-trained parameters and fine-tuned weights to correct spelling errors, grammatical inconsistencies, and structural noise while preserving the semantic content of the original consultation. No dialogue context or speaker information is incorporated.

### Full large language model based restoration

The Full LLM baseline transforms perturbed clinical consultation text into clinically corrected consultation text through a cascaded, instruction-driven processing workflow. Input texts with 10% or 100% perturbation are processed sequentially in batches to ensure manageable token lengths and maintain contextual integrity. Each batch undergoes a three-stage pipeline, combining structured instruction prompts with the LLM to perform content restoration. In the first stage, the LLM executes full-text content correction, removing noise, correcting fragmented phrases, and resolving transcription errors. Domain-specific features and general medical knowledge are integrated through prompts, allowing the model to preserve clinically relevant terminology while reconstructing coherent sentences. Non-clinical content such as hospital announcements or one-sided phone dialogues is filtered through embedded guidance in the instruction prompts. The second stage applies speaker-aware prompts, enabling the LLM to refine preliminary speaker labels and accurately distinguish doctors, patients, and other participants. The instruction specifies how content and speaker signals should be combined without additional classifier modules, leveraging the LLM’s understanding of dialogue context. In the final stage, a full-instruction consolidation is applied: the LLM integrates all prior corrections and speaker refinements into a unified, clinically coherent consultation text. Prompts guide the model to preserve medical accuracy, maintain linguistic fluency, and produce outputs that are immediately usable for downstream evaluation. The pipeline is implemented as a sequence of LLM calls, where each call receives the full set of relevant instructions for its subtask, avoiding iterative trial-and-error and minimizing redundant processing. The output from this cascaded instruction-driven pipeline serves as a baseline for semantic restoration evaluation. Evaluation metrics, including Accuracy, Integrity, and Smoothness, are applied in the same framework as PCRAgent to provide a direct point of comparison.

### Ethics and data access statement

Validation cohorts and multi-center clinical data used in this study were approved by the respective Institutional Review Boards of all participating sites (lead institution: [Hainan Medical University] [Harbin Medical University], approval no. [HYMLL-2026-109]). For the public discovery datasets (including AI Medical Chatbot dataset and MultiWOZ 2.2), ethical approval and informed consent had been obtained by the original data contributors; data access was strictly adhered to the data use policies of the respective repositories.

## Author contributions

M.Z., J.Z., W.T. and Y.Z. contributed to the conception and design of the study. M.Z., J.Z., W.T., J.L., J.X., H.Z. and J.Q. contributed to the data acquisition, curation and analysis. Y.Z. supervised the study. All authors contributed to the writing and revision of the manuscript and approved the final version.

## Data Availability

The public clinical dialogue corpus used in this study was obtained from the AI Medical Chatbot dataset, available at https://go.hyper.ai/W5OnS. The perturbed consultation sets for controlled noise evaluation were generated from this corpus according to the mixed level perturbation framework described in the Methods section. Real-world outpatient doctor patient dialogues used for multicenter clinical validation were collected from Harbin Medical University and Hainan Medical University. These data are not publicly available because they contain sensitive clinical information and are subject to patient privacy, ethical, and institutional data-use restrictions. Deidentified validation data may be available from the corresponding author upon reasonable request, subject to ethics approval and completion of an appropriate data use agreement.

## Code availability

The source code for PCRAgent is available on GitHub at https://github.com/TWjun1412/PCRAgent.

## Acknowledgments

This study was funded by Hainan Medical University Talent Research Start-up Fund (RZ2500003512). Undergraduate Research and Innovation Training Program of Hainan Medical University, (RZ2600001158)

