## Supplementary Figure S1-S4; Supplementary Table S1-S4 for "A Multi-Agent clinical pre-consultation system for structuring noisy patient reported information into clinical reports and AI-ready data"

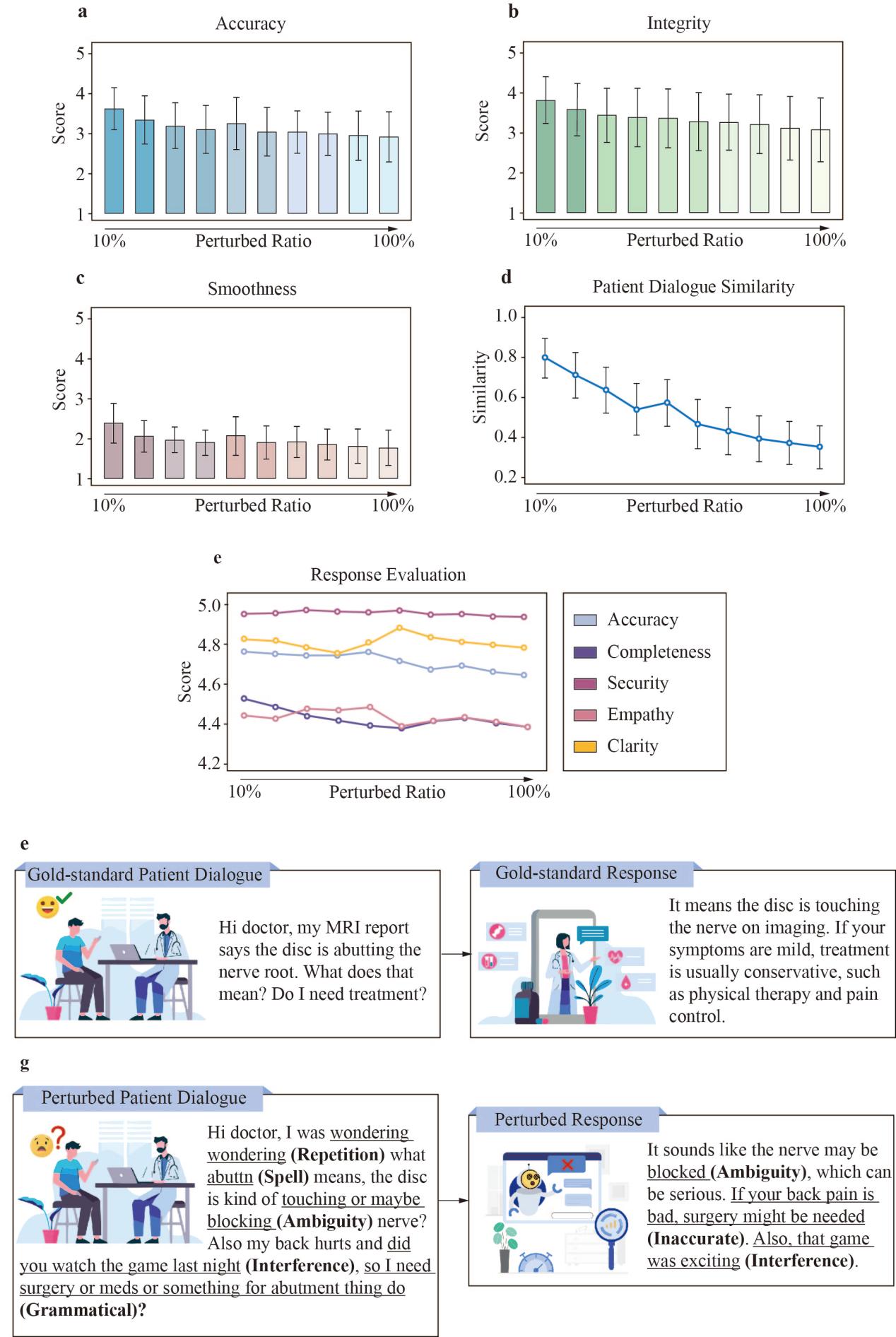

Figure S1. Impact of increasing noise on patient dialogue quality and downstream responses.

(a–d) Changes in patient dialogue-level metrics as the perturbation ratio increases from 0% to 100%, including Accuracy, Integrity, Smoothness, and Semantic Similarity. Each curve reflects GPT-4o’s direct processing of perturbed patient dialogues without semantic restoration, showing a monotonic degradation as noise intensifies. (e) Response-level evaluation scores generated from perturbed patient dialogues across Accuracy, Completeness, Security, Empathy, and Clarity, indicating how patient dialogue corruption propagates into downstream clinical recommendations. (f) An example of a Gold-standard patient dialogue and its corresponding response, illustrating coherent clinical intent and stable response quality under clean input conditions. (g) A representative perturbed patient dialogue and its generated response, highlighting typical failure patterns under noisy input, including disrupted semantics and degraded interaction quality.

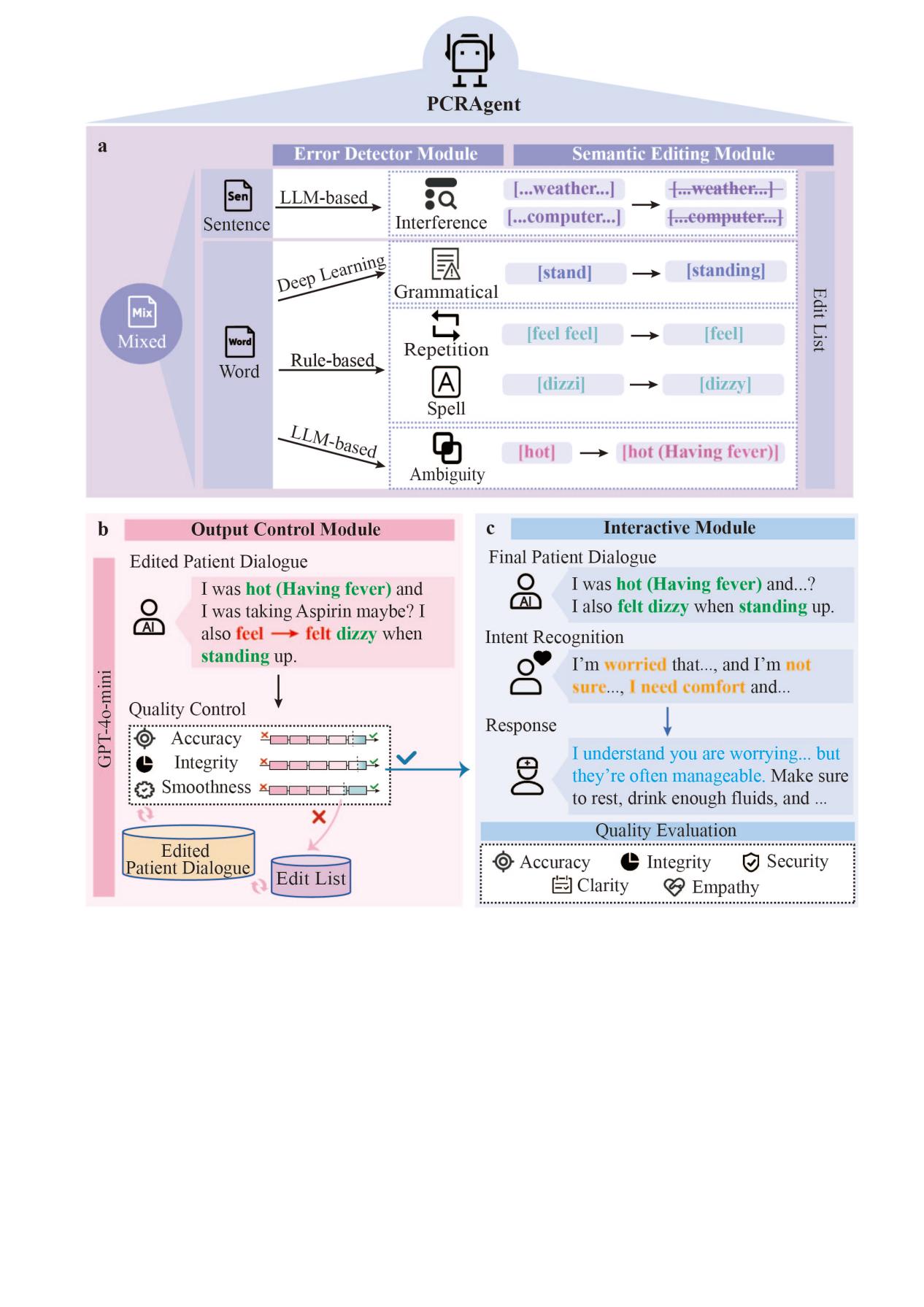

Figure S2. Internal coordination workflow of the PCRAgent framework.

(a) The Error Detector Module processes noisy patient dialogue inputs using a parallel feedback strategy across five error categories (spelling, repetitions, grammar, medical ambiguities and non-medical interference), while a medical knowledge graph protects valid clinical terms from being flagged as errors. The module outputs structured error annotations without rewriting the full text. The Semantic Editing Module then performs targeted restoration by editing only annotated regions, generating a traceable EditList that enables memory-based revisiting of unresolved issues in subsequent iterations. (b) The Output Control Module verifies the edited patient dialogue against three criteria (Accuracy, Integrity and Smoothness). Guided by the EditList, it selectively refines only non-compliant regions, iterating until all standards are met. (c) The Interactive Module performs anchor-guided analysis to interpret clinical semantics and patient intent.

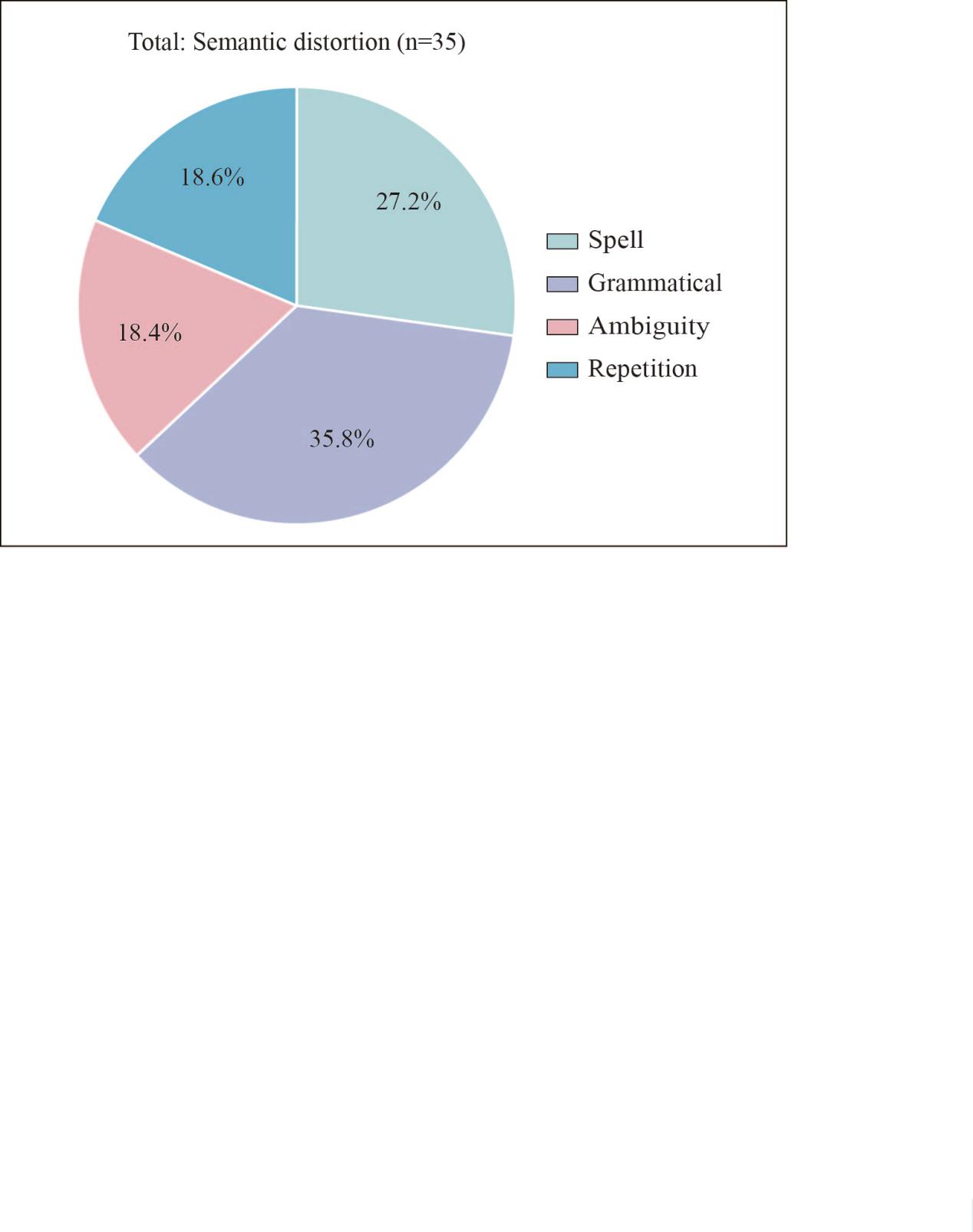

Figure S3. Distribution of error types observed in real medical communication.

Proportions of four common error types—spelling, grammatical inconsistency, ambiguity, and repetition—derived from manual transcription of 35 publicly available YouTube medical communication videos.

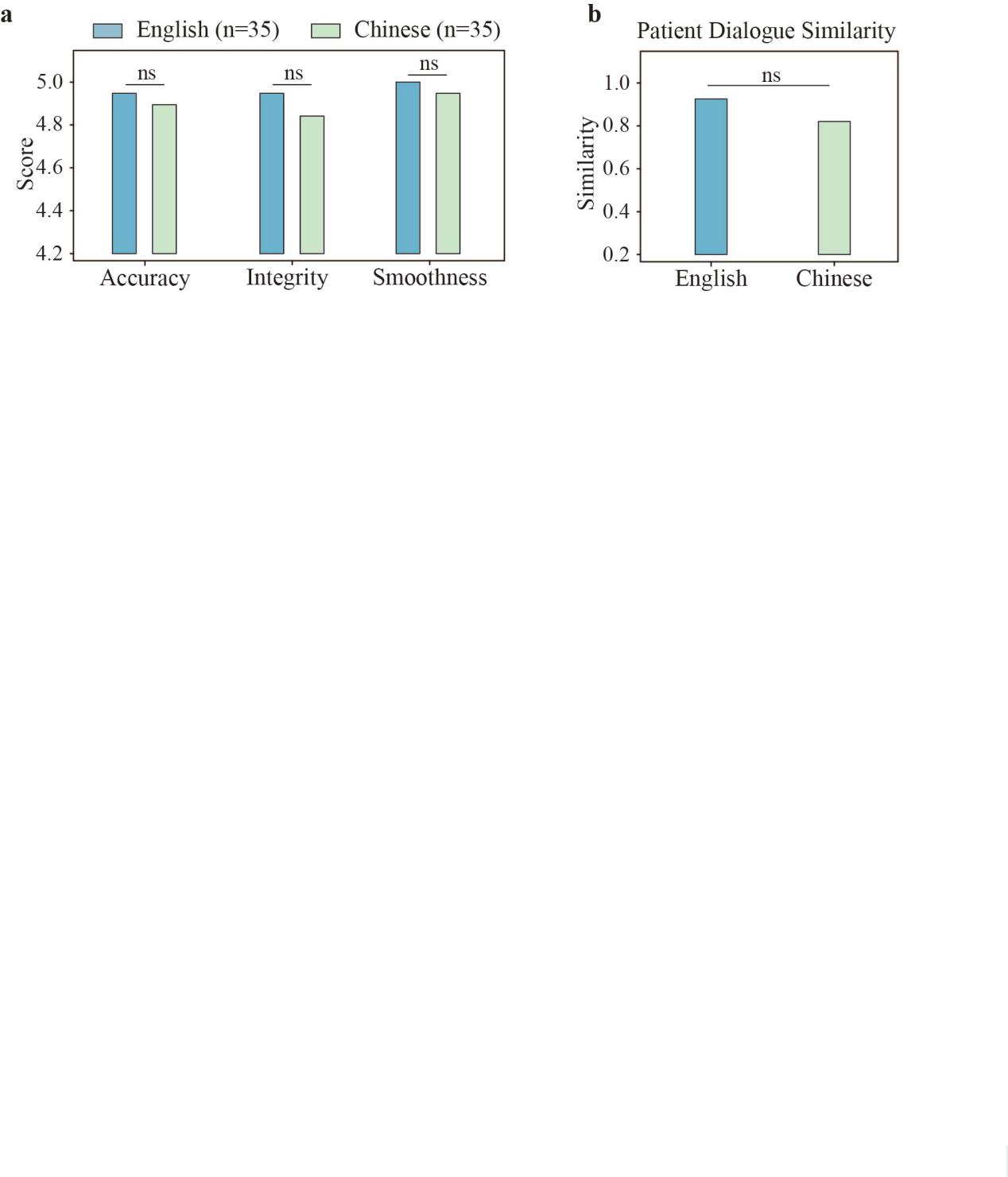

Figure S4. Cross-lingual robustness of PCRAgent under real-world clinical noise.

(a) Comparison of restoration quality across English and Chinese clinical patient dialogues evaluated on Accuracy, Integrity, and Smoothness. English patient dialogues are derived from real-world medical dialogue videos, while Chinese patient dialogues consist of authentic clinical cases. (b) Semantic Similarity between restored patient dialogues and reference text for English and Chinese inputs.

Table S1 Definition of the Edit Object in the EditList

| **Field** | **Type** | **Description** |
| --- | --- | --- |
| start_char | int | The starting character index of the edit span in the original patient dialogue text (0-based indexing). |
| end_char | int | The ending character index of the edit span (exclusive), defining the precise boundary of the affected region. |
| op | str | The edit operation type, including REPLACE, DELETE, or INSERT, specifying how the original text should be modified. |
| cand_texts | List[str] | A list of candidate replacement texts proposed by the detector or editing module. Multiple candidates indicate alternative correction hypotheses. |
| score | float | A confidence score in the range [0.0,1.0][0.0, 1.0][0.0,1.0], reflecting the detector’s reliability for this edit decision. |
| tag | str | The error category label associated with the edit: RPT (repetition), SPL (spelling), GRM (grammar), AMB (ambiguity), or ITF (non-medical interference). |
| edit_type | str | The edit generation mode: deterministic for rule-constrained edits, or candidate for edits requiring semantic selection. |
| detector_name | str | The name of the detector or tool that generated this edit, enabling provenance tracking across parallel detection modules. |

Table S2 Patient dialogue Restoration Quality Evaluation Metrics

| **Metric** | **Definition** | **Score = 1** | **Score = 3** | **Score = 5** |
| --- | --- | --- | --- | --- |
| **Accuracy** | Measures whether semantic restoration introduces hallucinated or factually incorrect medical content. | Introduces incorrect symptoms, conditions, or medical facts not present in the original patient dialogue. Negations or dosages are altered. | Minor factual ambiguity or slightly imprecise phrasing, but no explicit hallucinated medical content. | Fully faithful to the original patient dialogue. No hallucination, no factual deviation, and no polarity inversion. |
| **Integrity** | Assesses preservation of clinically essential information from the original patient dialogue. | Omits or distorts key clinical information such as symptoms, duration, medications, or test results. | Preserves main clinical facts but loses secondary details or contextual constraints. | All clinically relevant information is preserved without omission, distortion, or over-summarization. |
| **Smoothness** | Evaluates fluency and readability of the restored patient dialogue after editing. | Text is fragmented, repetitive, or difficult to read due to unnatural edits. | Generally readable with minor awkward phrasing or local incoherence. | Fluent, natural, and coherent, with edits seamlessly integrated into the original dialogue flow. |

Table S3 Recommendation Output Quality Evaluation Metrics

| **Metric** | **Definition** | **Score = 1** | **Score = 3** | **Score = 5** |
| --- | --- | --- | --- | --- |
| **Accuracy** | Verifies alignment of medical statements with established clinical knowledge. | Contains incorrect medical advice, hallucinated diagnoses, or unsupported clinical claims. | Mostly correct but includes vague or weakly supported medical statements. | Fully consistent with standard clinical knowledge and free from hallucinated content. |
| **Completeness** | Assesses coverage of clinically relevant aspects of the patient’s concerns. | Addresses only a small subset of the patient’s problems or ignores key concerns. | Covers main issues but misses secondary considerations or follow-up guidance. | Addresses all clinically relevant concerns with appropriate scope and depth. |
| **Security** | Screens for subtle Security risks in framing, reassurance, or interpretation. | Includes misleading reassurance, speculative interpretation, or advice that may delay care. | Generally safe but includes mildly ambiguous or weakly cautioned statements. | Explicitly avoids unsafe assumptions, provides appropriate caution, and maintains Security-oriented framing. |
| **Empathy** | Evaluates whether the response reflects inferred patient intent and emotional state. | Emotion is ignored or replaced by generic, template-based comforting language. | Shows basic emotional awareness but lacks intent-specific alignment. | Empathy is grounded in inferred patient intent and emotional cues derived from cleaned semantics. |
| **Clarity** | Measures whether the explanation and guidance are easy for patients to understand. | Disorganized, verbose, or confusing structure that hinders comprehension. | Understandable but with minor structural or expression issues. | Clearly structured, concise, and easy to follow for non-expert patients. |

Table S4 Examples of Intention Anchors in the Interactive Module

| **Emotional Category** | **Anchor Sentence (First-person, Explicit Form)** | **Typical Patient Surface Expression** | **Intended Response Strategy** |
| --- | --- | --- | --- |
| Seeking reassurance | *I am worried that my condition might be more serious than what has been said, and I need reassurance that it is under control.* | “Is this really nothing serious?” | Address concern directly before providing medical explanation. |
| Fear of deterioration | *I am afraid that my symptoms are getting worse and that the treatment may not be working.* | “The medicine doesn’t seem to help.” | Explain disease progression and clarify expected treatment response. |
| Doubt about diagnosis | *I am uncertain whether the diagnosis is correct and I am afraid of being misdiagnosed.* | “Are you sure this is the right diagnosis?” | Justify diagnosis with observable evidence and differential reasoning. |
| Medication anxiety | *I am worried about the side effects of the medication and how it might harm my body.* | “I’m a bit scared of taking this drug.” | Balance risk explanation with Security boundaries and monitoring advice. |
| Helplessness / loss of control | *I feel that I have little control over my condition and do not know what I should do next.* | “I don’t know what else I can do.” | Provide step-by-step guidance and emphasize actionable options. |
| Distrust in treatment | *I am unsure whether the prescribed treatment is necessary or effective.* | “Do I really need to take this?” | Explain treatment rationale and consequences of non-adherence. |
| Emotional overwhelm | *I feel emotionally overwhelmed by my symptoms and the uncertainty around them.* | “I’m really tired of all this.” | Acknowledge emotional burden before delivering clinical content. |
| Concern about long-term impact | *I am worried that this condition may affect my future life or daily functioning.* | “Will this affect me long-term?” | Discuss prognosis and realistic expectations without speculation. |
